# MAScINSIGHT: A Real-World Data Study to Deepen Knowledge and Management of McCune-Albright Syndrome – Study Protocol for an Observational Study within OHDSI

**DOI:** 10.64898/2026.10.24.26366390

**Authors:** Michele Zoch, Clemens Hentrich, Käty E. O. Mothes, Jens Weidner, Katja Hoffmann, Martin Sedlmayr, Angela Hübner, Christian Reich, Ines Reinecke, Georg Schwabe, Joline Wernsmann, Jens Bauer, Corinna Grasemann

## Abstract

**Introduction:** McCune-Albright syndrome (MAS) is a rare congenital disorder affecting endocrine, skeletal and dermatological systems. MAS is often coded as fibrous dysplasia (FD) in routine health data, which includes other conditions and does not allow precise identification. This coding gap complicates reliable epidemiological estimates and clinical characterization. The study MAScINSIGHT aims to address this by investigating MAS features within FD-coded cohorts.

**Methods and analysis:** MAScINSIGHT is a retrospective observational study using a federated network of data in the OMOP Common Data Model. Routine data from multiple sites will be analyzed within a federated setting to assess endocrine abnormalities and treatment patterns in patients with FD or MAS.

**Ethics and dissemination:** Analyses will be performed locally at the data partner, with only aggregate statistics made available and centrally analyzed. The study protocol was approved by the ethics committee (reference number: 2026-18760-retrospektiv). Findings will be disseminated through open-access publications and patient-oriented reports.

**Strengths and limitations of this study:**

- The study uses a federated approach based on the Observational Health Data Sciences and Informatics (OHDSI) framework and the OMOP Common Data Model, enabling standardized and reproducible analyses across institutions and countries while preserving data privacy.
- The multinational approach enables the analysis of larger and more diverse patient populations than typically available at individual institutions, addressing a major challenge in rare disease research.
- The integration of rare disease-specific terminologies, particularly ORPHAcodes, into the OHDSI vocabulary system supports more precise cohort identification and characterization, providing a transferable methodological approach for rare disease research.
- The retrospective design relies on routinely collected healthcare data, which may be affected by incomplete documentation, inconsistent coding practices and differences in data availability and quality across participating sites.
- Due to the rarity of McCune-Albright syndrome, sample sizes may remain limited despite the multinational approach, potentially restricting statistical power and subgroup analyses.

## Introduction

### 1.1 Rationale and background

McCune-Albright Syndrome (MAS) is a rare congenital mosaic disorder that affects the endocrine system, skeletal system and skin. The full clinical picture is referred to as Fibrous Dysplasia / McCune-Albright Syndrome (FD/MAS). It is caused by postzygotic activating mutations in the gene ‘guanine nucleotide binding protein, alpha stimulating activity polypeptide’ (GNAS), which encodes the Gs-alpha subunit of G protein-coupled receptors. Because the mutations are postzygotic, they lead to somatic mosaicism and clinical manifestations depend on the timing and location of the mutation during embryonic development [1,2].

MAS is defined by a triad of fibrous dysplasia (FD), café-au-lait skin macules and autonomous endocrine hyperfunction. While the condition is inborn, it usually manifests during childhood with variable age of onset and symptoms at onset. FD may be monostotic or polyostotic and frequently affects the craniofacial skeleton, leading to deformities, pain and neurovascular compressions that can result in blindness or hearing loss [3,4]. Endocrine features are marked by autonomous hormone production. Hyperthyroidism is frequently observed and presents with elevated free thyroxine (fT4) and suppressed levels of thyroid-stimulating hormone (TSH) [1,5]. Precocious puberty, particularly in girls, is characterized by elevated estradiol (E_2_) levels before the age of eight and suppressed luteinizing hormone (LH) levels due to the autonomous E_2_ production [6]. Growth hormone (GH) excess, though less common, is of particular concern due to its role in exacerbating craniofacial FD. Elevated Insulin-like Growth Factor 1 (IGF-1) values (> +2 standard deviation, adjusted for age and sex) suggest GH excess and should prompt further diagnostic work up [1,7]. Diagnosis is usually based on clinical criteria, often supported by biochemical markers. Genetic confirmation is possible in lesional tissue but is rarely necessary and remains challenging in peripheral blood due to absent or low levels of mosaicism [2].

Despite its distinctive phenotype, MAS is not specifically coded in the 10^th^ Revision of the International Statistical Classification of Diseases and Related Health Problems (ICD-10) coding system. Instead, it is grouped under the ICD-10 code for FD (Q78.1), which also includes the Jaffé-Lichtenstein Syndrome (JLS). While both conditions share FD, JLS is limited to the skeletal system and lacks endocrine or cutaneous involvement. This conflation of two distinct entities within the same code represents a major diagnostic and epidemiological challenge. However, MAS has its own ORPHAcode (562), which provides an opportunity for more specific case definition. The adoption of ORPHAcodes as an alternative terminology to identify unique rare diseases remains inconsistent [8,9]. This lack of specificity means that the true prevalence of MAS within the Q78.1-coded population is unknown. Patients with subclinical or partial manifestations of MAS may be misclassified or overlooked, esp. if endocrine abnormalities are not actively screened [3].

The coding ambiguity hinders identification in real-world data and limits epidemiological surveillance. A multinational retrospective analysis of harmonized routine data provides an opportunity to estimate the prevalence of MAS within the FD-coded population. Comparison with the ORPHAcode-coded cohort can help quantify the coding gap and provide insights into the clinical and therapeutic characteristics of MAS.

### 1.2 Study objectives

The primary objective is to investigate MAS features in FD patients in routine data from multiple international sites. The secondary objectives are to (1) assess the prevalence of endocrine abnormalities, (2) characterize treatment patterns and (3) evaluate the GH excess within both cohorts. Figure 1 illustrates the focus areas and corresponding research questions.

**Figure 1.**
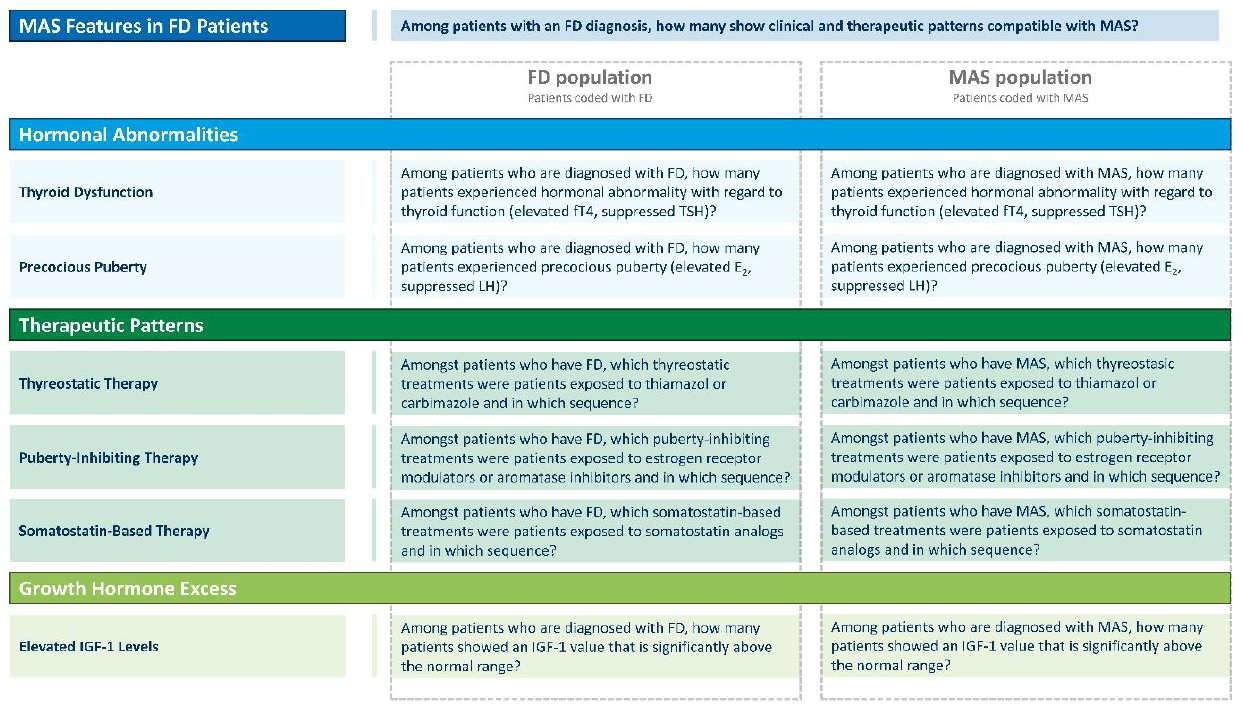
Thematic focus areas and corresponding research questions (E_2_: Estradiol, FD: Fibrous dysplasia, FSH: Follicle-stimulating hormone, fT4: Free thyroxine, IGF-1: Insulin-like Growth Factor 1, LH: Luteinizing hormone, MAS: McCune-Albright Syndrome, TSH: Thyroid-stimulating hormone)

## 2 Methods and analysis

### 2.1 Data sources

The study is based on routine data stored in OMOP Common Data Model (CDM). In the context of rare diseases, secondary data use present a great opportunity for evidence generation: It allows larger and more diverse study populations with less selection bias [10].

OMOP is an internationally established CDM maintained by Observational Health Data Sciences and Informatics (OHDSI). It is effective through its focus on relevant domains for observational data and of international terminologies by providing Standardized Vocabularies [11,12]. For the coding of rare diseases, ORPHAcodes are particularly useful for rare diseases [8]. Efforts are underway to integrate ORPHAcodes as an OMOP-conformant vocabulary [13]. Before their formal integration, the *Transition Database for Rare Diseases (TransDB)* is used, which consolidates mappings between ORPHAcodes and other terminologies such as ICD-10 and Systematized Nomenclature of Medicine (SNOMED) [14]. The table *SOURCE_TO_CONCEPT_MAP* can then be derived from the *TransDB* to use individual mappings within OMOP; the mapping of ORPHAcodes to SNOMED is freely accessible (https://doi.org/10.5281/zenodo.10817772).

### 2.2 Study process

The study will be conducted as a federated analysis: Data remain at the study sites and are analyzed locally; no patient-level data but only aggregate summary statistics are transmitted to the Study Coordination Center (SCC).

Local data are enriched using a centrally provided *SOURCE_TO_CONCEPT_MAP*, derived from the *TransDB*. Each site is responsible for its data transformation to the OMOP CDM, including quality assurance, supported by *ACHILLES HEEL* (https://ohdsi.github.io/Achilles/reference/achillesHeel.html) for identifying potential issues and the *Data Quality Dashboard* (*DQD*; https://github.com/OHDSI/DataQualityDashboard) for assessing completeness, conformance, and plausibility. Database characterization is performed with *ACHILLES* (descriptive summaries; https://github.com/OHDSI/Achilles) and *ATLAS* (definition of cohorts; https://github.com/ohdsi/atlas). Centrally developed R scripts are applied using *STRATEGUS* (https://github.com/OHDSI/Strategus) to coordinate the distributed workflow and *HADES* (https://github.com/OHDSI/Hades) package for data extraction, statistical modeling and calibration. Anonymized results are aggregated within a Shiny application. Figure 2 illustrates the different applications of the central and federated components. These components are then applied throughout an eight-step study process, based on distributed analyses of rare diseases that have already been conducted in Germany [9] (see Supplemental File A, Supplemental Figure 1).

**Figure 2.**
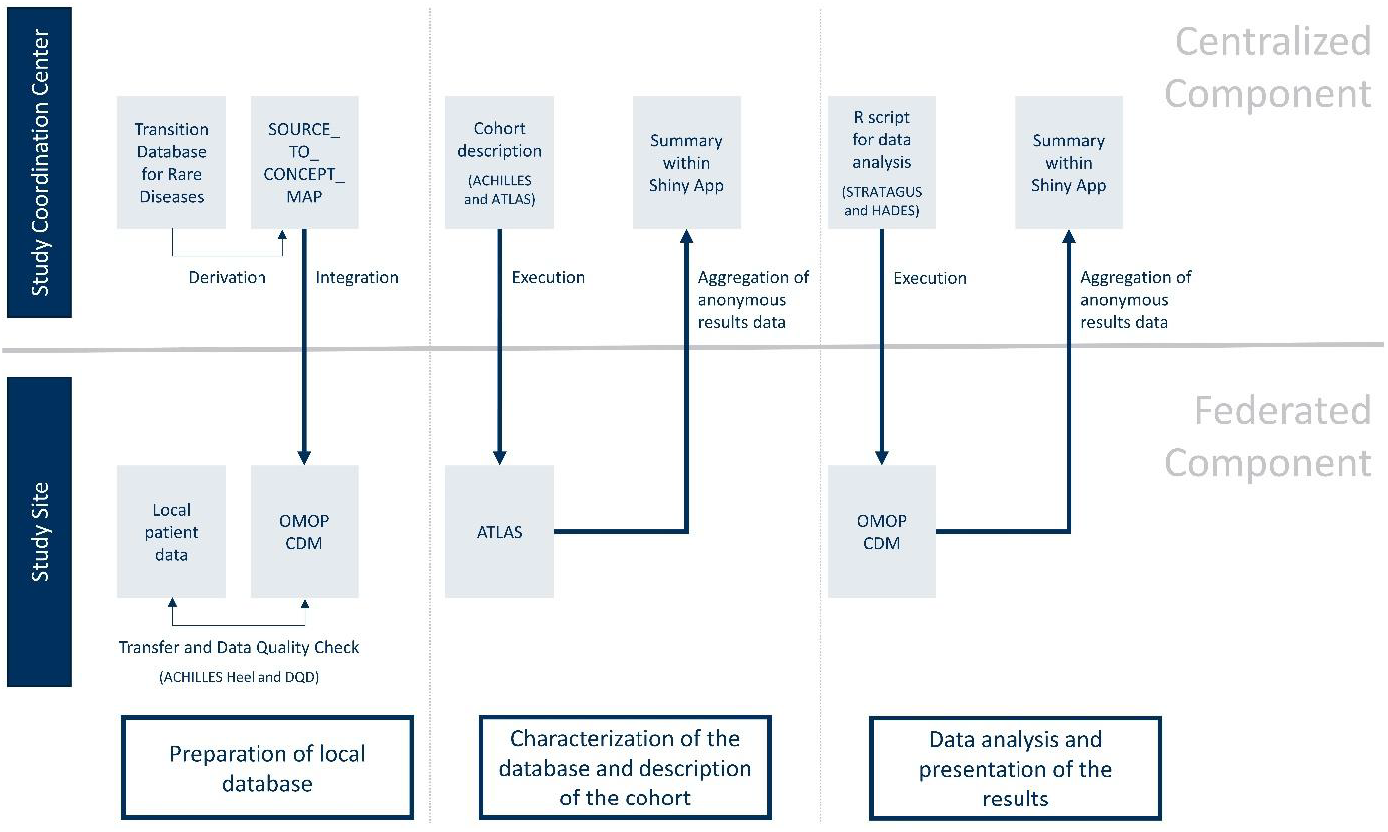
Centralized and federated components of the study

### 2.3 Outcome of interest

The primary outcome is the proportion of FD patients who meet MAS-criteria. Secondary outcomes include the prevalence of endocrine abnormalities, treatments and occurrence of GH excess. Figure 3 shows the focus areas, each linked to relevant medical concepts (see Supplemental File B, Supplemental Tables 1 – 8).

**Figure 3.**
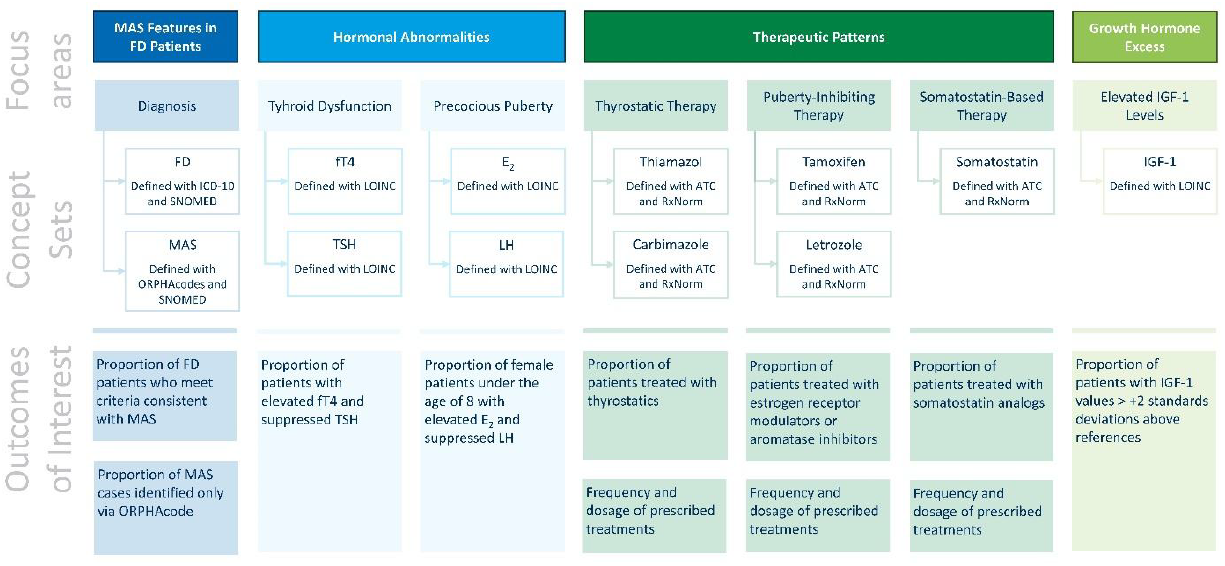
Conceptual mapping of concept sets and outcomes of interest (E_2_: Estradiol, FD: Fibrous dysplasia, fT4: Free thyroxine, IGF-1: Insulin-like Growth Factor 1, LH: Luteinizing hormone, MAS: McCune-Albright Syndrome, TSH: Thyroid-stimulating hormone)

### 2.4 Study population

The population comprises patients of all ages. For the primary research question, only a coded diagnosis is necessary; to answer the secondary research questions, data availability is also necessary (see Figure 4).

**Figure 4.**
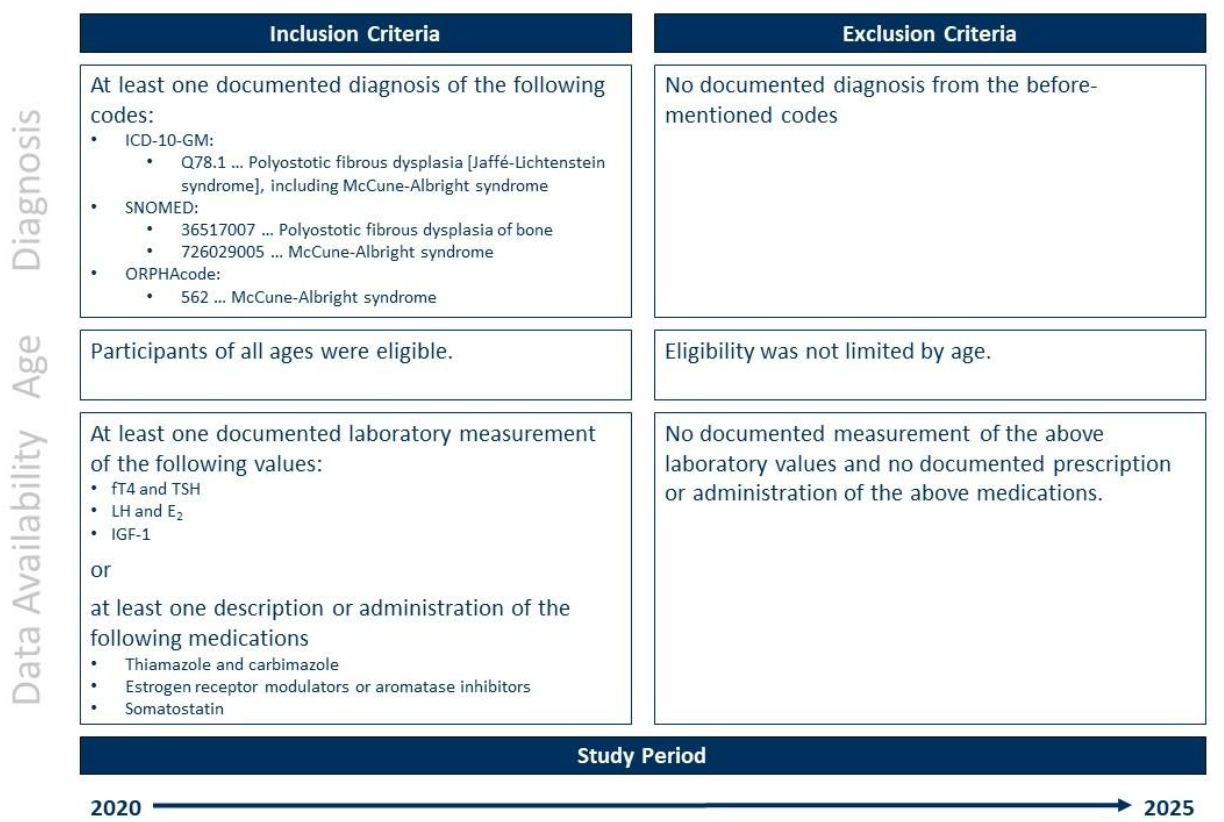
Characteristics of the study population, esp. inclusion and exclusion criteria as well as study period (data availability is only necessary for the secondary research questions)

Recruitment follows three phases:

1. Early data will be gathered from 2-3 sites to test feasibility of analyses scripts.
2. The national expansion will use research infrastructure of the German *Medical Informatics Initiative (MII)*.
3. The study will be extended to international level within *OHDSI*.

### 2.5 Statistical analysis

Clinical characteristics will be summarized using descriptive statistics. Group comparisons will apply χ^2^ or Fisher’s exact tests for categorical, and t- or Mann-Whitney U-tests for continuous variables. Effect sizes will be reported as odds ratios and risk ratios accompanied by 95% confidence intervals and mean differences or standardized mean differences. Subgroup analyses will be performed where appropriate. Confounding and bias will be addressed through propensity score weighting, negative controls and empirical calibration.

Analyses will be conducted using R and OHDSI tools. All scripts will be version-controlled and published in Git repositories.

### 2.6 Sample size and study power

The expected sample size for this study is anticipated to range from 80 – 120 patients. This estimate is based on published literature describing individual cases [5,7,15] as well as small series [6,16], with larger multicenter analyses including between 28 [17], 92 [18] and 113 patients [2]. In line with this, we expect to detect a comparable number of MAS cases within the FD population.

### 2.7 Strengths and limitations

The study is designed as a multi-site retrospective observational study based on secondary data use. Strength lies in reduced burden of data collection. By aggregating anonymized results, sensitive patient data remains securely stored within each site. Additional value is expected from the use of ORPHAcodes as an OMOP-conformant vocabulary for rare diseases, even prior to their integration into Standardized Vocabulary, to improve the identification of MAS.

Nonetheless, the risk of a Healthcare Process Bias must be acknowledged [9]: The routine data were originally documented for care-related and billing purposes, which may not reflect the true clinical status. Although the study promotes the use of ORPHAcodes, these are not yet mandatory across all sites.

## 3 Ethics and dissemination

### 3.1 Ethics

This study is conducted in strict accordance with ethical standards and regulatory frameworks. It exclusively makes use of secondary data, with these data being pseudonymized in OMOP locally at each site. The data transferred are aggregated results, which effectively eliminates re-identification of individual patients. Therefore, informed consent is not required.

The study follows the “Strengthening the Reporting of Observational Studies in Epidemiology” (STROBE) guidelines [19] (see Supplemental File C, Supplemental Table 9).

The study protocol has been reviewed and approved by the responsible ethics committee (reference number: 2026-18760-retrospektiv).

### 3.2 Dissemination

Peer-reviewed journal publications will present the study’s findings to the scientific community; patient-centered reports will translate them into accessible language. All analysis scripts, mapping tables and concept sets will be openly shared via Git repositories in line with Open Science principles. Aggregated results will be visualized through an interactive Shiny application.

## Data Availability

All data used to develop the study protocol are included in the manuscript or in its supplementary files.

## Declarations

### Author contributions

MZ was responsible for the technical and methodological conception of the study; CG was responsible for the medical conception of the study. IR, KH and CR provided support regarding the integration of the study into the OHDSI framework. JW provided support in the preparation of the statistical analysis plan. MZ led writing of the manuscript. All authors reviewed drafts and approved the final version.

### Ethics statements

Patient consent for publication: Not applicable.

Ethics approval: The Ethics Committee of the Landesärztekammer Rheinland Pfalz (reference number: 2026-18760-retrospektiv) has approved this observational study.

## Acknowledgements

This study is conducted in collaboration with the German patient advocacy group *FDMAS e. V*., which supports the overall research concept and contributes the patient perspective to the interpretation of the findings in light of patients’ lived experiences and healthcare realities, as well as to their patient-oriented dissemination.

The authors would like to express their gratitude to the *Service Center Research Data* of the TUD Dresden University of Technology for their valuable advice and support in the field of research data management; particularly for their constructive feedback during the preparation of the data management plan.

We gratefully acknowledge support from the SLUB / TU Dresden Open Access Publication Fund to enable the Open Access funding. The funders for the publication had no influence on the study design, data collection, analysis, decision to publish or preparation of the manuscript.

## Abbreviations

ATC: Anatomical Therapeutic Chemical
CDM: Common Data Model
DQD: Data Quality Dashboard
E_2_: Estradiol
FD: Fibrous dysplasia
FD / MAS: Fibrous Dysplasia / McCune-Albright Syndrome
fT3: Free thyrotropin
fT4: Free thyroxine
GH: Growth hormone
GNAS: Guanine nucleotide binding protein, alpha stimulating activity polypeptide
ICD-10: 10^th^ Revision of the International Statistical Classification of Diseases and Related Health Problems
IGF-1: Insulin-like Growth Factor 1
JLS: Jaffé-Lichtenstein Syndrome
LH: Luteinizing hormone
LOINC: Logical Observation Identifiers Names and Codes
MAS: McCune-Albright Syndrome
OHDSI: Observational Health Data Sciences and Informatics
SCC: Study Coordination Center
SNOMED: Systematized Nomenclature of Medicine
STROBE: Strengthening the reporting of observational studies in epidemiology
TransDB: Transition Database for Rare Diseases
TSH: Thyroid-stimulating hormone

