## Supplemental File A for "MAScINSIGHT: A Real-World Data Study to Deepen Knowledge and Management of McCune-Albright Syndrome – Study Protocol for an Observational Study within OHDSI"

Supplemental File A describes the process for conducting federated analyses within the MAScINSIGHT study. The study follows a federated research approach in which analytical scripts are centrally developed and executed locally at each participating site against harmonized data in the OMOP CDM. This framework ensures data privacy, reproducibility, and scalability across institutions, as only aggregated, summary statistics are shared for central analysis.

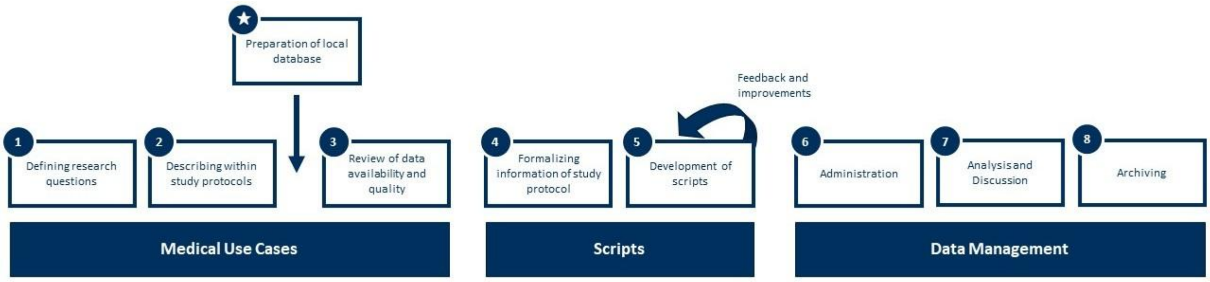


Supplemental Figure 1. Process for conducting federated analyses

Supplemental Figure 1 illustrates the federated analysis process, which is adapted from a previously published approach [9] and extended by an additional step "Preparation of local database". This step includes the enrichment of OMOP CDM instances with rare disease specific terminologies, such as the integration of ORPHAcodes via mapping approaches (e.g., TransDB), to enable consistent identification of rare disease cohorts.
