## Supplemental File B for "MAScINSIGHT: A Real-World Data Study to Deepen Knowledge and Management of McCune-Albright Syndrome – Study Protocol for an Observational Study within OHDSI"

Supplemental File B provides detailed definitions of all medical concepts used in the MAScINSIGHT study, including diagnoses, laboratory measurements and treatments. Concept definitions are based on standardized vocabularies within the OMOP CDM (e.g., SNOMED CT, LOINC, RxNorm) and, where necessary, extended through mappings of ORPHAcodes to SNOMED concepts. The use of standardized and reproducible concept sets ensures consistency of cohort definitions and comparability of results across participating sites.

#### Diagnoses

##### Fibrous dysplasia (FD)

Supplemental Table 1. Concept codes for fibrous dysplasia

| Medical concept | Terminology | Code | Description |
|---|---|---|---|
| FD | ICD-10-GM | Q78.1 | Polyostotic fibrous dysplasia [Jaffé-Lichtenstein syndrome]<br>Including McCune-Albright syndrome |
| FD | SNOMED | 76801 | Polyostotic fibrous dysplasia of bone |

##### McCune-Albright syndrome (MAS)

Supplemental Table 2. Concept codes for McCune-Alright syndrome

| Medical concept | Terminology | Code | Description |
|---|---|---|---|
| MAS | ORPHAnet | 562 | McCune-Albright syndrome |
| MAS | SNOMED | 726029005 | McCune-Albright syndrome |

### Hormonal abnormalities

#### Thyroid dysfunction

Supplemental Table 3. Concept codes for thyroid dysfunction

| Medical concept | Terminology | Code | Description |
|---|---|---|---|
| fT4 | LOINC | 58838-4 | Thyroxine (T4) free [Mass/volume] in Serum or Plasma – baseline |
| fT4 | LOINC | 27987-7 | Thyroxine (T4) free [Units/volume] in Serum or Plasma –baseline |
| fT4 | LOINC | 53349-7 | Thyroxine (T4) free [Mass/volume] in DBS |
| fT4 | LOINC | 53350-5 | Thyroxine (T4) free [Moles/volume] in DBS |
| fT4 | LOINC | 14920-3 | Thyroxine (T4) free [Moles/volume] in Serum or Plasma |
| fT4 | LOINC | 3024-7 | Thyroxine (T4) free [Mass/volume] in Serum or Plasma |
| fT4 | LOINC | 32215-6 | Thyroxine (T4) free index in Serum or Plasma by calculation |
| TSH | LOINC | 20452-9 | Thyrotropin [Presence] in Blood |
| TSH | LOINC | 48700-9 | Thyrotropin releasing hormone [Mass] of Dose |
| TSH | LOINC | 3015-5 | Thyrotropin [Units/volume] in Blood |
| TSH | LOINC | 35134-6 | Thyrotropin Ab [Presence] in Serum |
| TSH | LOINC | 3018-9 | Thyrotropin releasing hormone [Mass/volume] in Plasma |
| TSH | LOINC | 5384-3 | Thyrotropin Ab [Units/volume] in Serum |
| TSH | LOINC | 82659-4 | Thyrotropin receptor Ab [Presence] in Serum |
| TSH | LOINC | 3016-3 | Thyrotropin [Units/volume] in Serum or Plasma |
| TSH | LOINC | 14999-7 | Thyrotropin [Units/volume] in Serum or Plasma -- baseline |

#### Precocious puberty

Supplemental Table 4. Concept codes for precocious puberty

| Medical concept | Terminology | Code | Description |
|---|---|---|---|
| $E_2$ | LOINC | 13883-4 | Estradiol (E2).bioavailable/Estradiol (E2).total in Serum or Plasma |

| | | | |
|---|---|---|---|
| $E_2$ | LOINC | 14960-9 | Estradiol (E2) Free/Estradiol (E2).total in Serum or Plasma |
| $E_2$ | LOINC | 55857-7 | Estradiol (E2) [Mass/volume] in Body fluid |
| $E_2$ | LOINC | 55858-5 | Estradiol (E2) [Moles/volume] in Body fluid |
| $E_2$ | LOINC | 14715-7 | Estradiol (E2) [Moles/volume] in Serum or Plasma |
| $E_2$ | LOINC | 2243-4 | Estradiol (E2) [Mass/volume] in Serum or Plasma |
| LH | LOINC | 1599-0 | Lutropin [Mass/volume] in Serum or Plasma – baseline |
| LH | LOINC | 32328-7 | Lutropin [Units/volume] in Serum or Plasma -- baseline |
| LH | LOINC | 2579-1 | Lutropin [Moles/volume] in Serum or Plasma |
| LH | LOINC | 10501-5 | Lutropin [Units/volume] in Serum or Plasma |

### Therapeutic patterns

#### Thyrostatic therapy

Supplemental Table 5. Concept codes for thyrostatic therapy

| Medical concept | Terminology | Code | Description |
|---|---|---|---|
| Thiamazol | ATC | H03BB02 | Thiamazol |
| Thiamazol | ATC | H03BB52 | Thiamazol, Combinations |
| Methimazole | RxNorm | 6835 | Methimazole (Ingredient) |
| Carbimazol | ATC | H03BB01 | Carbimazol |
| Carbimazol | RxNorm | 2020 | Carbimazole (Ingredient) |

#### Puberty-inhibiting therapy

Supplemental Table 6. Concept codes for puberty-inhibiting therapy

| Medical concept | Terminology | Code | Description |
|---|---|---|---|
| Tamoxifen | ATC | L02BA01 | Tamoxifen |
| Tamoxifen | RxNorm | 10324 | Tamoxifen (Ingredient) |
| Letrozol | ATC | L02BG04 | Letrozol |
| Letrozol | RxNorm | 72965 | Letrozole (Ingredient) |

#### Somatostatin-based therapy

Supplemental Table 7. Concept codes for somatostatin-based therapy

| Medical concept | Terminology | Code | Description |
|---|---|---|---|
| Somatostatin | ATC | H01CB01 | Somatostatin |
| Somatostatin | RxNorm | 9939 | Somatostatin (Ingredient) |

### Growth hormone excess

#### Elevated IGF-1 levels

Supplemental Table 8. Concept codes for IGF-1

| Medical concept | Terminology | Code | Description |
|---|---|---|---|
| IGF-1 | LOINC | 32023-4 | Insulin-like growth factor-I [Mass/volume] in Blood |
| IGF-1 | LOINC | 13896-6 | Insulin-like growth factor-I [Units/volume] in Blood |
| IGF-1 | LOINC | 38492-5 | Insulin-like growth factor-I [Units/volume] in Serum or Plasma |
| IGF-1 | LOINC | 29775-4 | Insulin-like growth factor-I [Moles/volume] in Serum or Plasma |
| IGF-1 | LOINC | 2484-4 | Insulin-like growth factor-I [Mass/volume] in Serum or Plasma |
