## Supplemental File C for "MAScINSIGHT: A Real-World Data Study to Deepen Knowledge and Management of McCune-Albright Syndrome – Study Protocol for an Observational Study within OHDSI"

Supplemental File C presents the STOBE checklist of the MAScINSIGHT study. The checklist is provided to ensure transparent and complete reporting of this observational study and to facilitate critical appraisal and reproducibility. Each STROBE item is mapped to the corresponding section of the manuscript. Since this manuscript presents a study protocol for a retrospective observational study, no participant recruitment, data extraction or final analyses have been completed at the time of submission. Therefore, STROBE Items 13 – 17 (Results) are mapped as planned reporting rather than completed study findings.

Supplemental Table 9. Mapping of STROBE checklist items to the MASCINSIGHT study protocol

| Item Number | Item | Description | Corresponding section in manuscript |
|---|---|---|---|
| **Title and abstract** | | | |
| 1a | Title | The manuscript title identifies the study as MAScINSIGHT and describes it as a real-world data study focused on MAS within the OHDSI framework. The observational and protocol character of the study is further specified in the abstract. | Title |
| 1b | Abstract | The abstract summarizes the clinical background of MAS, the coding challenge within FD cohorts, the retrospective observational design using OMOP CDM, the federated analytical approach, and ethics and dissemination plans. As this is a study protocol, no study results are reported yet. | Abstract |
| **Introduction** | | | |
| 2 | Background / rationale | AAS is a rare multisystem disorder that is frequently coded under FD (ICD-10 Q78.1), which prevents precise identification in routine data. The study addresses this coding gap by using OMOP CDM, ORPHAcodes, and federated analyses to improve case identification and clinical characterization of MAS. | 1.1 Rationale and background |
| 3 | Objectives | The primary objective is to investigate MAS features in patients with FD using routinely collected data and to identify the proportion of FD patients who meet diagnostic criteria for MAS. Secondary objectives are to assess the prevalence of endocrine abnormalities, characterize treatment patterns, and evaluate growth hormone excess in both FD- and ORPHAcode-defined MAS cohorts | 1.2 Study objectives |
| **Methods** | | | |
| 4 | Study design | MAScINSIGHT is designed as a retrospective multicenter observational study using routinely collected healthcare data harmonized in the OMOP CDM. | 2.1 Data sources<br>2.2 Study process |

| | | | |
|---|---|---|---|
| | | Analyses are conducted in a federated setting, where data remain locally at participating sites and only anonymized aggregated results are centrally combined. The study follows a phased national and international expansion within the OHDSI framework. | |
| 5 | Setting | The study uses secondary routine care data from multiple national and international sites transformed into the OMOP CDM. Initial feasibility analyses will be conducted in 2 – 3 sites, followed by national expansion through the German MII and subsequent international collaboration within the OHDSI network. The study includes patients of all ages and uses retrospective data collected during routine clinical care, with site-specific observation periods depending on local data availability. | 2.1 Data sources<br>2.4 Study population |
| 6 | Participants | The study population includes patients of all ages with at least one documented diagnosis of FD or MAS identified through ICD-10, SNOMED or ORPHAcode concepts. For the primary research question, a coded diagnosis is sufficient for inclusion. For secondary analyses on endocrine abnormalities, treatment patterns, and growth hormone excess, additional availability of relevant laboratory values (e.g., fT4, TSH, LH, estradiol, IGF-1) or treatment records is required. Recruitment follows a phased approach from early feasibility testing to national and international expansion. | 2.4 Study population<br>Figure 4 |
| 7 | Variables | The primary outcome is the proportion of patients with FD who fulfill diagnostic criteria for MAS. Secondary variables include endocrine abnormalities (e.g., hyperthyroidism, precocious puberty, elevated IGF-1), treatment patterns (thyrostatic therapy, puberty-inhibiting therapy, somatostatin-based therapy), and occurrence of growth hormone excess. Diagnoses, laboratory measurements, medications and demographic characteristics are defined using standardized OMOP vocabularies including ICD-10, SNOMED, LOINC, ATC, RxNorm and ORPHAcodes. Detailed concept definitions and diagnostic criteria are provided in Appendix B. | 2.3 Outcome of interest<br>Appendix B |

| | | | |
|---|---|---|---|
| 8 | Data sources / measurement | Data are derived from routinely collected clinical care data stored locally at participating sites and harmonized in the OMOP CDM. Information on diagnoses, laboratory values, medications and demographic characteristics is extracted using standardized vocabularies such as ICD-10, SNOMED, LOINC, ATC, RxNorm and ORPHAcodes. ORPHAcodes are integrated through mappings to SNOMED using the TransDB and the SOURCE_TO_CONCEPT_MAP table. Data quality and comparability across sites are supported by ACHILLES HEEL, DQD, ACHILLES and ATLAS. Detailed medical concept definitions are provided in Appendix B. | 2.1 Data sources<br>Appendix B |
| 9 | Bias | Potential sources of bias include healthcare process bias, misclassification due to non-specific ICD-10 coding of MAS within FD, incomplete use of ORPHAcodes and variability in routine clinical documentation across sites. To reduce these risks, the study uses standardized OMOP vocabularies, ORPHAcode-to-SNOMED mappings via TransDB, federated analyses with harmonized scripts, data quality assessment tools (ACHILLES HEEL and DQD), propensity score weighting, negative controls, empirical calibration and subgroup analyses where appropriate. | 2.5 Statistical analysis<br>2.7 Strengths and limitations |
| 10 | Study size | The expected study size is approximately 80 – 120 patients. This estimate is based on published literature including individual case reports, small case series, and larger multicenter observational studies reporting between 28 and 113 patients with MAS. A comparable number of MAS cases is expected to be identified within the FD-coded population, providing sufficient feasibility for descriptive and comparative analyses. | 2.6 Sample size and study power |
| 11 | Quantitative variables | Quantitative variables include laboratory measurements such as fT4, TSH, LH, $E_2$ and IGF-1, as well as treatment dosages where available. Continuous variables will be summarized using means with standard deviations or medians with interquartile ranges, depending on distribution. Clinically relevant thresholds are applied for specific outcomes, such as elevated IGF-1 (> +2 standard deviations adjusted for age and sex) to indicate possible growth hormone excess and age-specific hormone abnormalities for precocious puberty and thyroid dysfunction. | 2.3 Outcome of interest<br>2.5 Statistical analysis |

| | | | |
|---|---|---|---|
| | | Categorization into clinically meaningful groups and subgroup analyses by age, sex and treatment periods are planned where appropriate. | |
| 12 | Statistical methods | Clinical characteristics will be summarized using descriptive statistics. Group comparisons will use $\chi^2$ or Fisher's exact tests for categorical variables and t-tests or Mann-Whitney U-tests for continuous variables. Effect sizes will be reported as odds ratios, risk ratios, mean differences, or standardized mean differences with 95% confidence intervals. Propensity score weighting, negative controls, and empirical calibration are planned to address confounding and systematic bias. Missing laboratory values may be handled by multiple imputation when appropriate. Subgroup analyses by age, sex, and treatment periods as well as sensitivity analyses are planned. If sufficient events are available, time-to-event analyses, mixed-effects models for site effects, and meta-analysis across sites may be performed. All analyses will be conducted using R and OHDSI tools, with scripts version-controlled and shared via Git repositories. | 2.5 Statistical analysis |
| **Results** | | | |
| 13 | Participants | Participant numbers will be reported for each stage of cohort selection, including patients with FD-coded diagnoses, patients with available laboratory or treatment data, patients fulfilling MAS criteria and final analytic cohorts. Reasons for exclusion due to missing data or unavailable relevant measurements will be documented where applicable. An aggregated federated cohort flow diagram is planned to visualize cohort derivation across participating sites. | 2.4 Study population<br>Figure 4 |
| 14 | Descriptive data | Demographic and clinical characteristics of study participants, including age, sex, diagnoses, endocrine abnormalities, laboratory values, treatments and relevant comorbidities, will be summarized descriptively for both FD-coded and ORPHAcode-defined MAS cohorts. Missing data for relevant laboratory variables will be reported where applicable. | 2.4 Study population<br>2.5 Statistical analysis |

| | | | |
|---|---|---|---|
| 15 | Outcome data | The primary outcome will be the proportion of patients with FD who fulfill diagnostic criteria for MAS. Secondary outcomes include frequencies of endocrine abnormalities, treatment patterns, medication dosages and occurrence of growth hormone excess. Outcome frequencies and summary measures will be reported separately for the relevant cohorts. | 2.3 Outcome of interest |
| 16 | Main results | Effect estimates for comparisons between cohorts, including odds ratios, risk ratios, mean differences and standardized mean differences with 95% confidence intervals, will be reported. Adjusted analyses using propensity score weighting and empirical calibration are planned where appropriate to address confounding and systematic bias. | 2.5 Statistical analysis |
| 17 | Other analyses | Planned additional analyses include subgroup analyses by age, sex and treatment periods, sensitivity analyses for missing data and coding assumptions, and, where feasible, time-to-event analyses, mixed-effects models for site effects and meta-analyses across participating sites to assess heterogeneity and robustness of findings. | 2.5 Statistical analysis |
| **Discussion** | | | |
| 18 | Key results | The study aims to quantify the coding gap between FD and MAS in routine data and to improve identification of MAS cases using OMOP CDM, ORPHAcodes and federated analyses. Key planned outcomes include prevalence estimates of endocrine abnormalities, characterization of treatment patterns and evaluation of growth hormone excess within FD- and MAS-defined cohorts. | 1.2 Study objectives<br>2.7 Strengths and limitations |
| 19 | Limitations | Potential limitations include healthcare process bias due to the use of routine care data, incomplete or inconsistent coding of MAS within ICD-10, variable adoption of ORPHAcodes across sites and possible misclassification caused by differences in local documentation practices. Additional limitations arise from retrospective data collection and the absence of prospective validation. These risks are addressed through standardized vocabularies, data quality tools and federated analytical methods. | 2.7 Strengths and limitations |

| | | | |
|---|---|---|---|
| 20 | Interpretation | The study is designed to improve epidemiological understanding and clinical characterization of MAS by addressing coding ambiguity and enabling standardized analyses across institutions and countries. The integration of ORPHAcodes into the OHDSI framework is expected to strengthen case identification, support more reliable real-world evidence generation and contribute to evidence-based care pathways for rare endocrine disorders. | 2.7 Strengths and limitations<br>3 Ethics and dissemination |
| 21 | Generalizability | The multinational and federated study design supports broad applicability of findings across different healthcare settings by using standardized OMOP vocabularies and harmonized OHDSI tools. Inclusion of national and international sites improves representativeness and enables transferability of methods and findings to other rare diseases facing similar coding and interoperability challenges. | 2.7 Strengths and limitations |
| **Other information** | | | |
| 22 | Funding | The study received no specific grant from any funding agency in the public, commercial or not-for-profit sectors. The manuscript explicitly states that no external funding was provided for the research. | Declarations |
